# Prevalence and Risk Factors of Respiratory Tract Infections Following Medically-Attended-Diarrhea in Children Aged 6-35 Months: Enterics for Global Health (EFGH)-*Shigella* Surveillance Study, 2022-2024

**DOI:** 10.64898/2026.04.17.26351078

**Authors:** Bakary Conteh, Sean R. Galagan, Henry Badji, Ousman Secka, Beth Tippett Bar, Sonia I. Rao, Hannah Atlas, Richard Omore, John B. Ochieng, Milagritos Tapia, Jen Cornick, Nigel Cunliffe, Loyda Fiorella Zegarra Paredes, Josh Colston, Md Taufiqul Islam, Md. Parvej Mosharraf, Farah Naz Qamar, Irum Fatima, Patricia B. Pavlinac, M. Jahangir Hossain

## Abstract

Globally, respiratory tract infections (RTI) are the main cause of morbidity, and in Low-middle-income countries (LMICs) RTI including pneumonia are a leading cause of morbidity and mortality in children <5 years. Diarrheal illness increases RTI risk in young children through micronutrient depletion, and immune stress, yet data on post-diarrhea RTI burden in LMICs are limited. We determined the prevalence and risk factors of RTI within three months following medically-attended diarrhea (MAD) in children aged 6-35 months enrolled in seven EFGH country sites in Asia, Africa and South America. The EFGH study prospectively enrolled children aged 6-35 months with MAD in selected health facilities during a 24-month period from 2022 to 2024 and followed them for three months. RTI was defined as cough or difficulty breathing and the presence of one of the following symptoms at any scheduled or unscheduled visit during follow-up: stridor; fast-breathing; oxygen saturation <90%; or chest indrawing. The period prevalence and 95% confidence intervals of RTI were calculated, and correlates of RTI were assessed using modified-Poisson regression. From June 2022 to August 2024, 9,476 children aged 6-35 months presenting with MAD in the EFGH study sites were screened: 9,116 (96.2%) included in the current study. Nearly half were female (46.7%), and median age was 15 months. Overall, 48.5% received all age-appropriate vaccines, and 87.6% received the pneumococcal vaccine, with significant variation across countries. Nearly one-quarter of children were stunted, 17.2% wasted, and 21.9% underweight. RTI occurred in 3.8% of children during the three-month follow-up, mostly within the first month. Higher prevalence of RTI occurred among children aged 12-23 months (8.7%), those undernourished (16.1%), unvaccinated (4.0%) or living in poor sanitation settings (4.1%). While children who received all age-appropriate or pneumococcal vaccinations had a lower crude prevalence of RTI, these associations were not statistically significant after adjusting for age, sex and study site. RTI was infrequently observed in the three months following MAD presentation, with significant variability by site and with the highest prevalence in Malawi. RTI risk was highest in 12-23-month-olds and among children with undernutrition, and those living in poor sanitation conditions.

## Introduction

Respiratory tract infections (RTIs) are a leading cause of morbidity and mortality in children under 5 years of age, especially in LMICs located in sub-Saharan Africa and South East Asia - the same region where approximately 80% of the annual global child diarrhea deaths continue to occur.(1–8) Low socioeconomic status is associated with pneumonia morbidity.(9) Large family size, low maternal education, inadequate water and sanitation systems, and household crowding are environmental factors that promote the transmission of respiratory pathogens.(1,3,5,10–12) Diarrheal disease is the third leading cause of death in children.(13) Children in LMICs settings suffering from frequent diarrhea episodes are also at a high risk of pneumonia.(10) Diarrhea is strongly associated with malnutrition which has multiple deleterious effects likely impacting RTI recovery.(14) Comorbid RTIs and diarrhea can also result in worse outcomes and children presenting to care with MAD may potentially benefit from pneumonia interventions.(10) Therefore, characterizing the burden and risk factors of RTI in children with diarrhea, a population likely at high risk of morbidity and mortality, is critical. Although there are existing interventions to reduce the burden of non-diarrheal illnesses, interventions that target children at particular risk may be most efficient and cost-effective. Diarrhea increases the risk of RTI and thus may be an ideal intervention point, particularly diarrhea that has led to care-seeking, for interventions that prevent future RTI.

The objective of this secondary data analysis is to describe the period prevalence and risk factors of RTIs in the three months following MAD in children aged 6-35 months in the study population of the Enterics for Global Health *shigella* surveillance study, which was conducted in Bangladesh, Pakistan, The Gambia, Malawi, Mali, Kenya and Peru.

## Methods Study design

This was a secondary analysis of data originally collected from an observational, prospective cohort study among children presenting with MAD to the EFGH-Shigella surveillance sentinel health facilities.

### Setting

This study was conducted in the seven EFGH country sites in Africa, Asia, and Latin America (Peru, Pakistan, Bangladesh, Mali, Malawi, Kenya, and The Gambia). A full description of EFGH Shigella surveillance study sites has been published previously.(15)

### Participants

We used complete-case analysis approach in this secondary analysis. We included all children with MAD 6-35 months of age enrolled into the EFGH study having completed scheduled follow up visits at either one month or three months or both. Additional information was collected from participants who returned to the facility during unscheduled visits. This entailed caregiver interview, clinical history recording and anthropometric measurements.

### Ethical consideration

This study was approved by the Institutional Review Boards of each EFGH country site. Informed consent was obtained from each participant before enrolment.

### Statistical analysis

The primary outcome was RTI during follow-up, defined as cough or difficulty breathing and the presence of one of the following symptoms at any visit during follow-up: (1) stridor; (2) fast-breathing (respiratory rate ≥50 breaths/minute for children less than one year of age and ≥40 breaths/minute for children one year or older); (3) oxygen saturation <90%; or (4) chest indrawing. Potential correlates of RTI of interest included age, sex, vaccination history, malnutrition, diarrhea severity, the presence of children under five years of age in the household, water, sanitation and hygiene (WASH) access as defined by The WHO/UNICEF Joint Monitoring Programme for Water Supply, Sanitation and Hygiene (JMP), socioeconomic status, and caregiver education.

The period prevalence of RTI was described by study timepoint at four week and three month follow up, and during unscheduled visits, stratified by age group (6–11, 12–17, 18–23 and 24–35 months), and stratified by potential correlates of interest. Additionally, the underlying symptoms (cough, difficulty breathing, stridor, oxygen saturation <90%, chest indrawing and fast breathing) and alternate definitions of RTIs (caregiver report, clinician diagnosis) were presented for the entire study period and stratified by study time-point. Finally, modified-Poisson regression with an exchangeable correlation matrix was used to determine the unadjusted and adjusted (for EFGH site, age category and sex) correlates of RTI infection. All analyses were conducted in R (version 4.5.0).

## Results

From June 2022 to August 2024, 9,476 children aged 6-35 months with an acute episode of MAD were enrolled in EFGH study; 9,116 (96.2%) were included in the current study (Fig 1). The most common reason for exclusion from analysis included the participant being lost to follow-up following enrollment (139/9476 [1.5%]) and withdrawing from the study (117/9476 [1.2%]) (Fig 1). Among the 9,116 children included in the analysis, there was a relatively even distribution of children enrolled from each of the seven country sites (Table 1). Children 6 –17 months of age had the highest number of participants enrolled (62.3%) and the median age was 14 months (interquartile range [IQR]: 09–21). Nearly half of enrolled children were female (45.7%). Approximately half of the children had received all age-appropriate vaccines (48.5%) according to national Expanded Programme on Immunization (EPI) vaccine schedules, with variability between countries; Bangladesh had the highest coverage (77.3%) and the Gambia the lowest (24.8%). Pneumococcal vaccine coverage was 85.8% with variability by site (from 58.1% in The Gambia to 98.1% in Kenya). The majority of mothers of enrolled children (67.9%) had completed primary school education or higher. Nearly one-quarter (23.5%) of enrolled children were stunted, 17.1% were wasted, and 21.9% were underweight at enrollment.

**Table 1.**
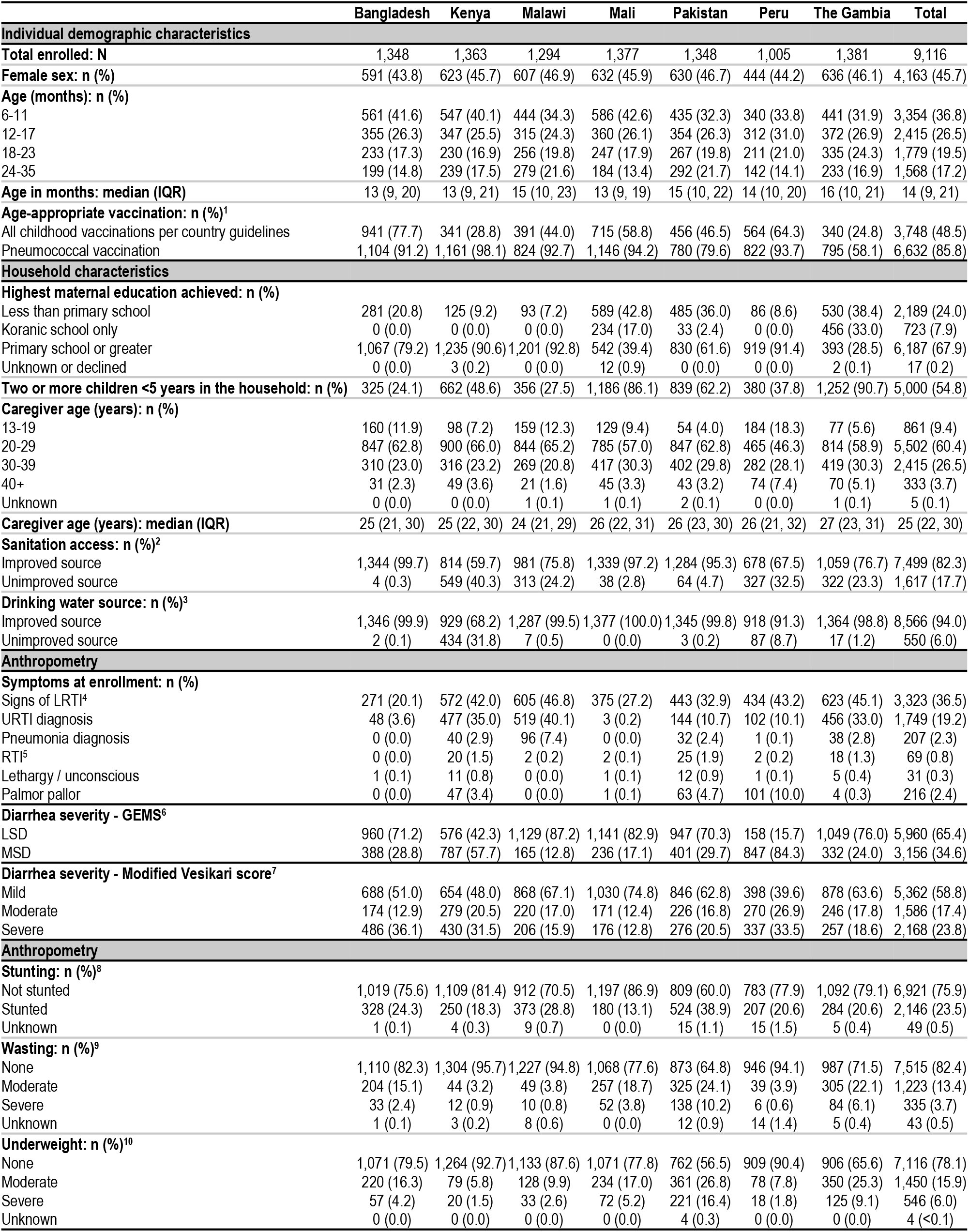
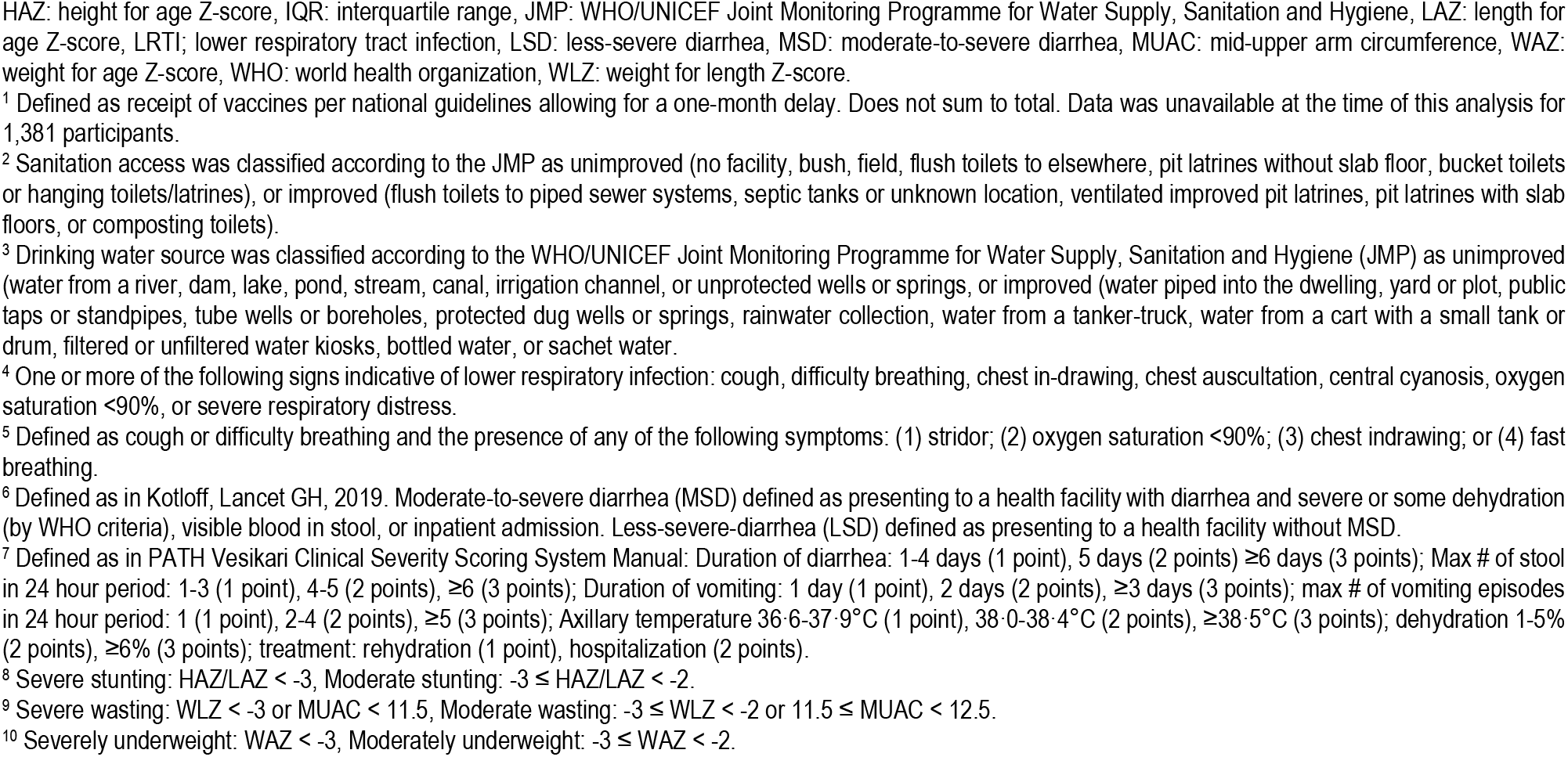
Demographics of study participants in the analysis set.

|  | Bangladesh | Kenya | Malawi | Mali | Pakistan | Peru | The Gambia | Total |
| --- | --- | --- | --- | --- | --- | --- | --- | --- |
| **Individual demographic characteristics** |  |  |  |  |  |  |  |  |
| Total enrolled: N | 1,348 | 1,363 | 1,294 | 1,377 | 1,348 | 1,005 | 1,381 | 9,116 |
| Female sex: n (%) | 591 (43.8) | 623 (45.7) | 607 (46.9) | 632 (45.9) | 630 (46.7) | 444 (44.2) | 636 (46.1) | 4,163 (45.7) |
| Age (months): n (%) |  |  |  |  |  |  |  |  |
| 6-11 | 561 (41.6) | 547 (40.1) | 444 (34.3) | 586 (42.6) | 435 (32.3) | 340 (33.8) | 441 (31.9) | 3,354 (36.8) |
| 12-17 | 355 (26.3) | 347 (25.5) | 315 (24.3) | 360 (26.1) | 354 (26.3) | 312 (31.0) | 372 (26.9) | 2,415 (26.5) |
| 18-23 | 233 (17.3) | 230 (16.9) | 256 (19.8) | 247 (17.9) | 267 (19.8) | 211 (21.0) | 335 (24.3) | 1,779 (19.5) |
| 24-35 | 199 (14.8) | 239 (17.5) | 279 (21.6) | 184 (13.4) | 292 (21.7) | 142 (14.1) | 233 (16.9) | 1,568 (17.2) |
| Age in months: median (IQR) | 13 (9, 20) | 13 (9, 21) | 15 (10, 23) | 13 (9, 19) | 15 (10, 22) | 14 (10, 20) | 16 (10, 21) | 14 (9, 21) |
| Age-appropriate vaccination: n (%)<sup>1</sup> |  |  |  |  |  |  |  |  |
| All childhood vaccinations per country guidelines | 941 (77.7) | 341 (28.8) | 391 (44.0) | 715 (58.8) | 456 (46.5) | 564 (64.3) | 340 (24.8) | 3,748 (48.5) |
| Pneumococcal vaccination | 1,104 (91.2) | 1,161 (98.1) | 824 (92.7) | 1,146 (94.2) | 780 (79.6) | 822 (93.7) | 795 (58.1) | 6,632 (85.8) |
| **Household characteristics** |  |  |  |  |  |  |  |  |
| Highest maternal education achieved: n (%) |  |  |  |  |  |  |  |  |
| Less than primary school | 281 (20.8) | 125 (9.2) | 93 (7.2) | 589 (42.8) | 485 (36.0) | 86 (8.6) | 530 (38.4) | 2,189 (24.0) |
| Koranic school only | 0 (0.0) | 0 (0.0) | 0 (0.0) | 234 (17.0) | 33 (2.4) | 0 (0.0) | 456 (33.0) | 723 (7.9) |
| Primary school or greater | 1,067 (79.2) | 1,235 (90.6) | 1,201 (92.8) | 542 (39.4) | 830 (61.6) | 919 (91.4) | 393 (28.5) | 6,187 (67.9) |
| Unknown or declined | 0 (0.0) | 3 (0.2) | 0 (0.0) | 12 (0.9) | 0 (0.0) | 0 (0.0) | 2 (0.1) | 17 (0.2) |
| Two or more children <5 years in the household: n (%) | 325 (24.1) | 662 (48.6) | 356 (27.5) | 1,186 (86.1) | 839 (62.2) | 380 (37.8) | 1,252 (90.7) | 5,000 (54.8) |
| Caregiver age (years): n (%) |  |  |  |  |  |  |  |  |
| 13-19 | 160 (11.9) | 98 (7.2) | 159 (12.3) | 129 (9.4) | 54 (4.0) | 184 (18.3) | 77 (5.6) | 861 (9.4) |
| 20-29 | 847 (62.8) | 900 (66.0) | 844 (65.2) | 785 (57.0) | 847 (62.8) | 465 (46.3) | 814 (58.9) | 5,502 (60.4) |
| 30-39 | 310 (23.0) | 316 (23.2) | 269 (20.8) | 417 (30.3) | 402 (29.8) | 282 (28.1) | 419 (30.3) | 2,415 (26.5) |
| 40+ | 31 (2.3) | 49 (3.6) | 21 (1.6) | 45 (3.3) | 43 (3.2) | 74 (7.4) | 70 (5.1) | 333 (3.7) |
| Unknown | 0 (0.0) | 0 (0.0) | 1 (0.1) | 1 (0.1) | 2 (0.1) | 0 (0.0) | 1 (0.1) | 5 (0.1) |
| Caregiver age (years): median (IQR) | 25 (21, 30) | 25 (22, 30) | 24 (21, 29) | 26 (22, 31) | 26 (23, 30) | 26 (21, 32) | 27 (23, 31) | 25 (22, 30) |
| Sanitation access: n (%)<sup>2</sup> |  |  |  |  |  |  |  |  |
| Improved source | 1,344 (99.7) | 814 (59.7) | 981 (75.8) | 1,339 (97.2) | 1,284 (95.3) | 678 (67.5) | 1,059 (76.7) | 7,499 (82.3) |
| Unimproved source | 4 (0.3) | 549 (40.3) | 313 (24.2) | 38 (2.8) | 64 (4.7) | 327 (32.5) | 322 (23.3) | 1,617 (17.7) |
| Drinking water source: n (%)<sup>3</sup> |  |  |  |  |  |  |  |  |
| Improved source | 1,346 (99.9) | 929 (68.2) | 1,287 (99.5) | 1,377 (100.0) | 1,345 (99.8) | 918 (91.3) | 1,364 (98.8) | 8,566 (94.0) |
| Unimproved source | 2 (0.1) | 434 (31.8) | 7 (0.5) | 0 (0.0) | 3 (0.2) | 87 (8.7) | 17 (1.2) | 550 (6.0) |
| **Anthropometry** |  |  |  |  |  |  |  |  |
| Symptoms at enrollment: n (%) |  |  |  |  |  |  |  |  |
| Signs of LRTI<sup>4</sup> | 271 (20.1) | 572 (42.0) | 605 (46.8) | 375 (27.2) | 443 (32.9) | 434 (43.2) | 623 (45.1) | 3,323 (36.5) |
| URTI diagnosis | 48 (3.6) | 477 (35.0) | 519 (40.1) | 3 (0.2) | 144 (10.7) | 102 (10.1) | 456 (33.0) | 1,749 (19.2) |
| Pneumonia diagnosis | 0 (0.0) | 40 (2.9) | 96 (7.4) | 0 (0.0) | 32 (2.4) | 1 (0.1) | 38 (2.8) | 207 (2.3) |
| RTI<sup>5</sup> | 0 (0.0) | 20 (1.5) | 2 (0.2) | 2 (0.1) | 25 (1.9) | 2 (0.2) | 18 (1.3) | 69 (0.8) |
| Lethargy / unconscious | 1 (0.1) | 11 (0.8) | 0 (0.0) | 1 (0.1) | 12 (0.9) | 1 (0.1) | 5 (0.4) | 31 (0.3) |
| Palmar pallor | 0 (0.0) | 47 (3.4) | 0 (0.0) | 1 (0.1) | 63 (4.7) | 101 (10.0) | 4 (0.3) | 216 (2.4) |
| Diarrhea severity - GEMS<sup>6</sup> |  |  |  |  |  |  |  |  |
| LSD | 960 (71.2) | 576 (42.3) | 1,129 (87.2) | 1,141 (82.9) | 947 (70.3) | 158 (15.7) | 1,049 (76.0) | 5,960 (65.4) |
| MSD | 388 (28.8) | 787 (57.7) | 165 (12.8) | 236 (17.1) | 401 (29.7) | 847 (84.3) | 332 (24.0) | 3,156 (34.6) |
| Diarrhea severity - Modified Vesikari score<sup>7</sup> |  |  |  |  |  |  |  |  |
| Mild | 688 (51.0) | 654 (48.0) | 868 (67.1) | 1,030 (74.8) | 846 (62.8) | 398 (39.6) | 878 (63.6) | 5,362 (58.8) |
| Moderate | 174 (12.9) | 279 (20.5) | 220 (17.0) | 171 (12.4) | 226 (16.8) | 270 (26.9) | 246 (17.8) | 1,586 (17.4) |
| Severe | 486 (36.1) | 430 (31.5) | 206 (15.9) | 176 (12.8) | 276 (20.5) | 337 (33.5) | 257 (18.6) | 2,168 (23.8) |
| **Anthropometry** |  |  |  |  |  |  |  |  |
| Stunting: n (%)<sup>8</sup> |  |  |  |  |  |  |  |  |
| Not stunted | 1,019 (75.6) | 1,109 (81.4) | 912 (70.5) | 1,197 (86.9) | 809 (60.0) | 783 (77.9) | 1,092 (79.1) | 6,921 (75.9) |
| Stunted | 328 (24.3) | 250 (18.3) | 373 (28.8) | 180 (13.1) | 524 (38.9) | 207 (20.6) | 284 (20.6) | 2,146 (23.5) |
| Unknown | 1 (0.1) | 4 (0.3) | 9 (0.7) | 0 (0.0) | 15 (1.1) | 15 (1.5) | 5 (0.4) | 49 (0.5) |
| Wasting: n (%)<sup>9</sup> |  |  |  |  |  |  |  |  |
| None | 1,110 (82.3) | 1,304 (95.7) | 1,227 (94.8) | 1,068 (77.6) | 873 (64.8) | 946 (94.1) | 987 (71.5) | 7,515 (82.4) |
| Moderate | 204 (15.1) | 44 (3.2) | 49 (3.8) | 257 (18.7) | 325 (24.1) | 39 (3.9) | 305 (22.1) | 1,223 (13.4) |
| Severe | 33 (2.4) | 12 (0.9) | 10 (0.8) | 52 (3.8) | 138 (10.2) | 6 (0.6) | 84 (6.1) | 335 (3.7) |
| Unknown | 1 (0.1) | 3 (0.2) | 8 (0.6) | 0 (0.0) | 12 (0.9) | 14 (1.4) | 5 (0.4) | 43 (0.5) |
| Underweight: n (%)<sup>10</sup> |  |  |  |  |  |  |  |  |
| None | 1,071 (79.5) | 1,264 (92.7) | 1,133 (87.6) | 1,071 (77.8) | 762 (56.5) | 909 (90.4) | 906 (65.6) | 7,116 (78.1) |
| Moderate | 220 (16.3) | 79 (5.8) | 128 (9.9) | 234 (17.0) | 361 (26.8) | 78 (7.8) | 350 (25.3) | 1,450 (15.9) |
| Severe | 57 (4.2) | 20 (1.5) | 33 (2.6) | 72 (5.2) | 221 (16.4) | 18 (1.8) | 125 (9.1) | 546 (6.0) |
| Unknown | 0 (0.0) | 0 (0.0) | 0 (0.0) | 0 (0.0) | 4 (0.3) | 0 (0.0) | 0 (0.0) | 4 (<0.1) |

HAZ: height for age Z-score, IQR: interquartile range, JMP: WHO/UNICEF Joint Monitoring Programme for Water Supply, Sanitation and Hygiene, LAZ: length for age Z-score, LRTI: lower respiratory tract infection, LSD: less-severe diarrhea, MSD: moderate-to-severe diarrhea, MUAC: mid-upper arm circumference, WAZ: weight for age Z-score, WHO: world health organization, WLZ: weight for length Z-score.
<sup>1</sup> Defined as receipt of vaccines per national guidelines allowing for a one-month delay. Does not sum to total. Data was unavailable at the time of this analysis for 1,381 participants.
<sup>2</sup> Sanitation access was classified according to the JMP as unimproved (no facility, bush, field, flush toilets to elsewhere, pit latrines without slab floor, bucket toilets or hanging toilets/latrines), or improved (flush toilets to piped sewer systems, septic tanks or unknown location, ventilated improved pit latrines, pit latrines with slab floors, or composting toilets).
<sup>3</sup> Drinking water source was classified according to the WHO/UNICEF Joint Monitoring Programme for Water Supply, Sanitation and Hygiene (JMP) as unimproved (water from a river, dam, lake, pond, stream, canal, irrigation channel, or unprotected wells or springs, or improved (water piped into the dwelling, yard or plot, public taps or standpipes, tube wells or boreholes, protected dug wells or springs, rainwater collection, water from a tanker-truck, water from a cart with a small tank or drum, filtered or unfiltered water kiosks, bottled water, or sachet water).
<sup>4</sup> One or more of the following signs indicative of lower respiratory infection: cough, difficulty breathing, chest in-drawing, chest auscultation, central cyanosis, oxygen saturation <90%, or severe respiratory distress.

**Fig 1.**
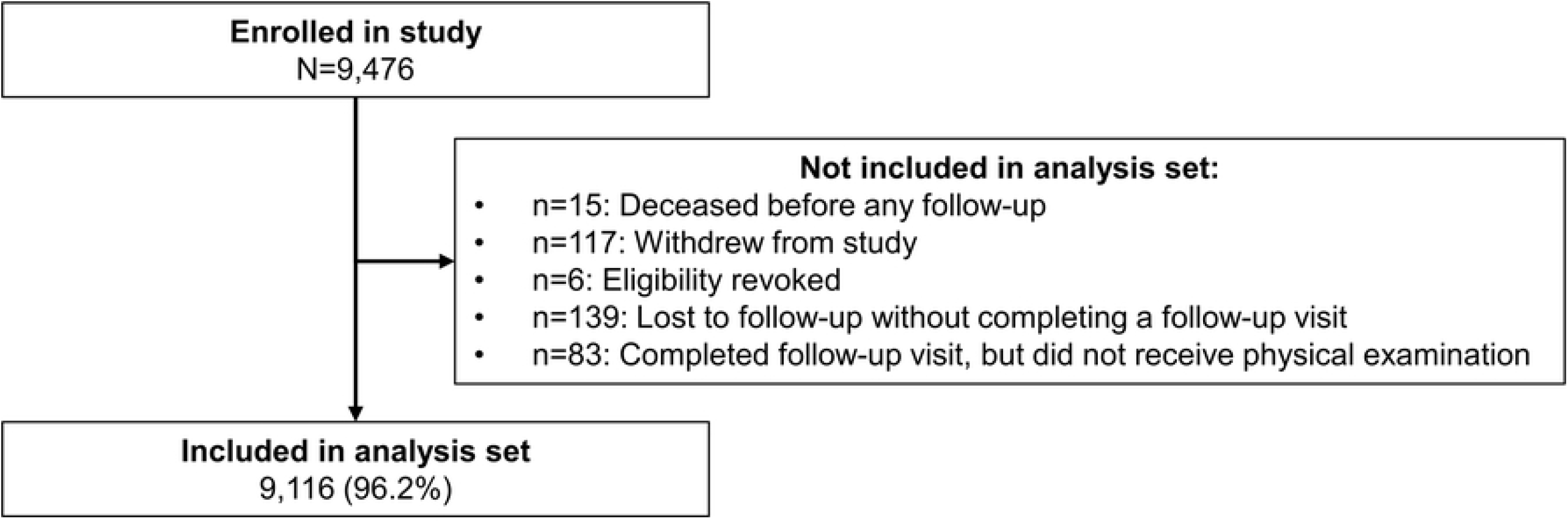
EFGH participant enrolment and inclusion in secondary analysis.

Respiratory tract infection occurred in 344 (3.8%) children in the three-month follow-up period (Table 2), most commonly in Malawi (13.8%) and least frequently in Mali (0.4%). Cough was the most common component of the RTI definition followed by fast breathing. RTI peaked in children aged 12– 17 months of age (4.4%) and was similar in children 18–23 months of age (4.3%) and 24–35 months of age (4.3%) and appeared lowest (2.8%) in children aged 6-11 months (Fig 2). Children who had received all age-appropriate vaccinations and pneumococcal vaccination had lower period prevalence of RTI (2.4% and 3.0%, respectively) compared to those who had not received all age-appropriate and pneumococcal vaccination (4.0% and 4.4%, respectively) (Table 3). RTI was prevalent in children with stunting (4.4%), wasting (5.2%), underweight (6.7%) and where there is poor sanitation (4.1%).

**Table 2.** Prevalence of RTI and its components study country site.

| Outcome | Bangladesh | Kenya | Malawi | Mali | Pakistan | Peru | The Gambia | Total |
| --- | --- | --- | --- | --- | --- | --- | --- | --- |
| **N** | 1,348 | 1,363 | 1,294 | 1,377 | 1,348 | 1,005 | 1,381 | 9,116 |
| **Clinical symptoms: n (%)** |  |  |  |  |  |  |  |  |
| Cough | 400 (29.7) | 240 (17.6) | 755 (58.3) | 57 (4.1) | 414 (30.7) | 24 (2.4) | 285 (20.6) | 2,175 (23.9) |
| Difficulty breathing | 6 (0.4) | 9 (0.7) | 31 (2.4) | 3 (0.2) | 17 (1.3) | 3 (0.3) | 15 (1.1) | 84 (0.9) |
| Stridor | 0 (0.0) | 1 (0.1) | 5 (0.4) | 0 (0.0) | 1 (0.1) | 0 (0.0) | 7 (0.5) | 14 (0.2) |
| Oxygen saturation <90% | 0 (0.0) | 2 (0.1) | 1 (0.1) | 0 (0.0) | 16 (1.2) | 0 (0.0) | 0 (0.0) | 19 (0.2) |
| Chest indrawing | 6 (0.4) | 0 (0.0) | 9 (0.7) | 0 (0.0) | 18 (1.3) | 1 (0.1) | 12 (0.9) | 46 (0.5) |
| Fast breathing<sup>1</sup> | 10 (0.7) | 49 (3.6) | 255 (19.7) | 10 (0.7) | 102 (7.6) | 7 (0.7) | 57 (4.1) | 490 (5.4) |
| RTI<sup>2</sup> | 7 (0.5) | 23 (1.7) | 179 (13.8) | 5 (0.4) | 78 (5.8) | 3 (0.3) | 49 (3.5) | 344 (3.8) |

<sup>1</sup> Defined as a respiratory rate of 50+ breaths/minute for children less than one year of age and 40+ for children one year of age or older.
<sup>2</sup> Primary outcome defined as cough or difficulty breathing and the presence of any of the following symptoms: (1) stridor; (2) oxygen saturation <90%; (3) chest indrawing; or (4) fast breathing.

**Table 3.**
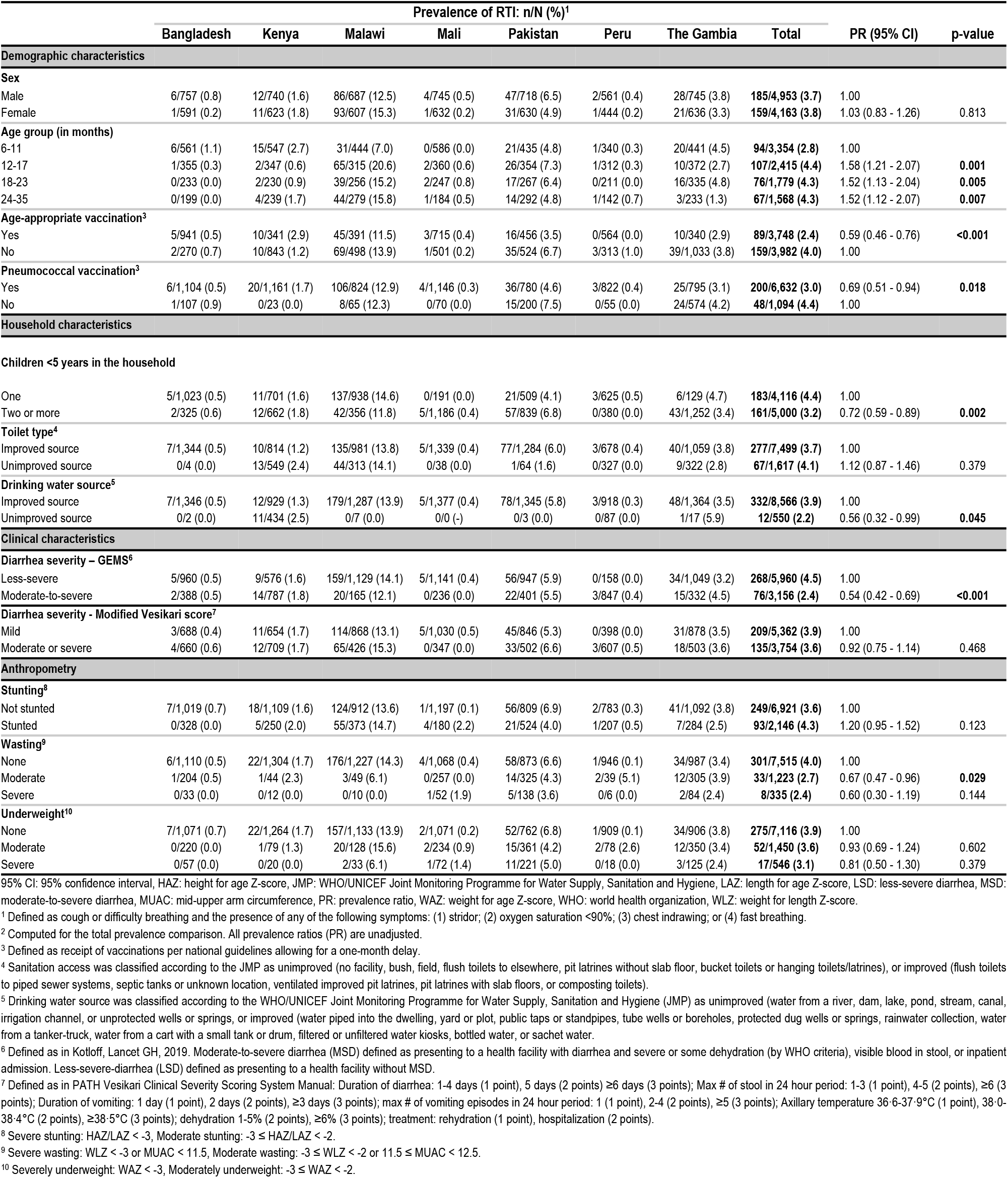
Prevalence of RTI by EFGH country site and demographics and clinical characteristics.

|  | Prevalence of RTI: n/N (%)<sup>1</sup> |  |  |  |  |  |  |  |  |  |
| --- | --- | --- | --- | --- | --- | --- | --- | --- | --- | --- |
|  | Bangladesh | Kenya | Malawi | Mali | Pakistan | Peru | The Gambia | Total | PR (95% CI) | p-value |
| Demographic characteristics |  |  |  |  |  |  |  |  |  |  |
| Sex |  |  |  |  |  |  |  |  |  |  |
| Male | 6/757 (0.8) | 12/740 (1.6) | 86/687 (12.5) | 4/745 (0.5) | 47/718 (6.5) | 2/561 (0.4) | 28/745 (3.8) | 185/4,953 (3.7) | 1.00 |  |
| Female | 1/591 (0.2) | 11/623 (1.8) | 93/607 (15.3) | 1/632 (0.2) | 31/630 (4.9) | 1/444 (0.2) | 21/636 (3.3) | 159/4,163 (3.8) | 1.03 (0.83 - 1.26) | 0.813 |
| Age group (in months) |  |  |  |  |  |  |  |  |  |  |
| 6-11 | 6/561 (1.1) | 15/547 (2.7) | 31/444 (7.0) | 0/586 (0.0) | 21/435 (4.8) | 1/340 (0.3) | 20/441 (4.5) | 94/3,354 (2.8) | 1.00 |  |
| 12-17 | 1/355 (0.3) | 2/347 (0.6) | 65/315 (20.6) | 2/360 (0.6) | 26/354 (7.3) | 1/312 (0.3) | 10/372 (2.7) | 107/2,415 (4.4) | 1.58 (1.21 - 2.07) | 0.001 |
| 18-23 | 0/233 (0.0) | 2/230 (0.9) | 39/256 (15.2) | 2/247 (0.8) | 17/267 (6.4) | 0/211 (0.0) | 16/335 (4.8) | 76/1,779 (4.3) | 1.52 (1.13 - 2.04) | 0.005 |
| 24-35 | 0/199 (0.0) | 4/239 (1.7) | 44/279 (15.8) | 1/184 (0.5) | 14/292 (4.8) | 1/142 (0.7) | 3/233 (1.3) | 67/1,568 (4.3) | 1.52 (1.12 - 2.07) | 0.007 |
| Age-appropriate vaccination<sup>3</sup> |  |  |  |  |  |  |  |  |  |  |
| Yes | 5/941 (0.5) | 10/341 (2.9) | 45/391 (11.5) | 3/715 (0.4) | 16/456 (3.5) | 0/564 (0.0) | 10/340 (2.9) | 89/3,748 (2.4) | 0.59 (0.46 - 0.76) | <0.001 |
| No | 2/270 (0.7) | 10/843 (1.2) | 69/498 (13.9) | 1/501 (0.2) | 35/524 (6.7) | 3/313 (1.0) | 39/1,033 (3.8) | 159/3,982 (4.0) | 1.00 |  |
| Pneumococcal vaccination<sup>3</sup> |  |  |  |  |  |  |  |  |  |  |
| Yes | 6/1,104 (0.5) | 20/1,161 (1.7) | 106/824 (12.9) | 4/1,146 (0.3) | 36/780 (4.6) | 3/822 (0.4) | 25/795 (3.1) | 200/6,632 (3.0) | 0.69 (0.51 - 0.94) | 0.018 |
| No | 1/107 (0.9) | 0/23 (0.0) | 8/65 (12.3) | 0/70 (0.0) | 15/200 (7.5) | 0/55 (0.0) | 24/574 (4.2) | 48/1,094 (4.4) | 1.00 |  |
| Household characteristics |  |  |  |  |  |  |  |  |  |  |
| Children <5 years in the household |  |  |  |  |  |  |  |  |  |  |
| One | 5/1,023 (0.5) | 11/701 (1.6) | 137/938 (14.6) | 0/191 (0.0) | 21/509 (4.1) | 3/625 (0.5) | 6/129 (4.7) | 183/4,116 (4.4) | 1.00 |  |
| Two or more | 2/325 (0.6) | 12/662 (1.8) | 42/356 (11.8) | 5/1,186 (0.4) | 57/839 (6.8) | 0/380 (0.0) | 43/1,252 (3.4) | 161/5,000 (3.2) | 0.72 (0.59 - 0.89) | 0.002 |
| Toilet type<sup>4</sup> |  |  |  |  |  |  |  |  |  |  |
| Improved source | 7/1,344 (0.5) | 10/814 (1.2) | 135/981 (13.8) | 5/1,339 (0.4) | 77/1,284 (6.0) | 3/678 (0.4) | 40/1,059 (3.8) | 277/7,499 (3.7) | 1.00 |  |
| Unimproved source | 0/4 (0.0) | 13/549 (2.4) | 44/313 (14.1) | 0/38 (0.0) | 1/64 (1.6) | 0/327 (0.0) | 9/322 (2.8) | 67/1,617 (4.1) | 1.12 (0.87 - 1.46) | 0.379 |
| Drinking water source<sup>5</sup> |  |  |  |  |  |  |  |  |  |  |
| Improved source | 7/1,346 (0.5) | 12/929 (1.3) | 179/1,287 (13.9) | 5/1,377 (0.4) | 78/1,345 (5.8) | 3/918 (0.3) | 48/1,364 (3.5) | 332/8,566 (3.9) | 1.00 |  |
| Unimproved source | 0/2 (0.0) | 11/434 (2.5) | 0/7 (0.0) | 0/0 (-) | 0/3 (0.0) | 0/87 (0.0) | 1/17 (5.9) | 12/550 (2.2) | 0.56 (0.32 - 0.99) | 0.045 |
| Clinical characteristics |  |  |  |  |  |  |  |  |  |  |
| Diarrhea severity – GEMS<sup>6</sup> |  |  |  |  |  |  |  |  |  |  |
| Less-severe | 5/960 (0.5) | 9/576 (1.6) | 159/1,129 (14.1) | 5/1,141 (0.4) | 56/947 (5.9) | 0/158 (0.0) | 34/1,049 (3.2) | 268/5,960 (4.5) | 1.00 |  |
| Moderate-to-severe | 2/388 (0.5) | 14/787 (1.8) | 20/165 (12.1) | 0/236 (0.0) | 22/401 (5.5) | 3/847 (0.4) | 15/332 (4.5) | 76/3,156 (2.4) | 0.54 (0.42 - 0.69) | <0.001 |
| Diarrhea severity - Modified Vesikari score<sup>7</sup> |  |  |  |  |  |  |  |  |  |  |
| Mild | 3/688 (0.4) | 11/654 (1.7) | 114/868 (13.1) | 5/1,030 (0.5) | 45/846 (5.3) | 0/398 (0.0) | 31/878 (3.5) | 209/5,362 (3.9) | 1.00 |  |
| Moderate or severe | 4/660 (0.6) | 12/709 (1.7) | 65/426 (15.3) | 0/347 (0.0) | 33/502 (6.6) | 3/607 (0.5) | 18/503 (3.6) | 135/3,754 (3.6) | 0.92 (0.75 - 1.14) | 0.468 |
| Anthropometry |  |  |  |  |  |  |  |  |  |  |
| Stunting<sup>8</sup> |  |  |  |  |  |  |  |  |  |  |
| Not stunted | 7/1,019 (0.7) | 18/1,109 (1.6) | 124/912 (13.6) | 1/1,197 (0.1) | 56/809 (6.9) | 2/783 (0.3) | 41/1,092 (3.8) | 249/6,921 (3.6) | 1.00 |  |
| Stunted | 0/328 (0.0) | 5/250 (2.0) | 55/373 (14.7) | 4/180 (2.2) | 21/524 (4.0) | 1/207 (0.5) | 7/284 (2.5) | 93/2,146 (4.3) | 1.20 (0.95 - 1.52) | 0.123 |
| Wasting<sup>9</sup> |  |  |  |  |  |  |  |  |  |  |
| None | 6/1,110 (0.5) | 22/1,304 (1.7) | 176/1,227 (14.3) | 4/1,068 (0.4) | 58/873 (6.6) | 1/946 (0.1) | 34/987 (3.4) | 301/7,515 (4.0) | 1.00 |  |
| Moderate | 1/204 (0.5) | 1/44 (2.3) | 3/49 (6.1) | 0/257 (0.0) | 14/325 (4.3) | 2/39 (5.1) | 12/305 (3.9) | 33/1,223 (2.7) | 0.67 (0.47 - 0.96) | 0.029 |
| Severe | 0/33 (0.0) | 0/12 (0.0) | 0/10 (0.0) | 1/52 (1.9) | 5/138 (3.6) | 0/6 (0.0) | 2/84 (2.4) | 8/335 (2.4) | 0.60 (0.30 - 1.19) | 0.144 |
| Underweight<sup>10</sup> |  |  |  |  |  |  |  |  |  |  |
| None | 7/1,071 (0.7) | 22/1,264 (1.7) | 157/1,133 (13.9) | 2/1,071 (0.2) | 52/762 (6.8) | 1/909 (0.1) | 34/906 (3.8) | 275/7,116 (3.9) | 1.00 |  |
| Moderate | 0/220 (0.0) | 1/79 (1.3) | 20/128 (15.6) | 2/234 (0.9) | 15/361 (4.2) | 2/78 (2.6) | 12/350 (3.4) | 52/1,450 (3.6) | 0.93 (0.69 - 1.24) | 0.602 |
| Severe | 0/57 (0.0) | 0/20 (0.0) | 2/33 (6.1) | 1/72 (1.4) | 11/221 (5.0) | 0/18 (0.0) | 3/125 (2.4) | 17/546 (3.1) | 0.81 (0.50 - 1.30) | 0.379 |

95% CI: 95% confidence interval, HAZ: height for age Z-score, JMP: WHO/UNICEF Joint Monitoring Programme for Water Supply, Sanitation and Hygiene, LAZ: length for age Z-score, LSD: less-severe diarrhea, MSD: moderate-to-severe diarrhea, MUAC: mid-upper arm circumference, PR: prevalence ratio, WAZ: weight for age Z-score, WHO: world health organization, WLZ: weight for length Z-score.
<sup>1</sup> Defined as cough or difficulty breathing and the presence of any of the following symptoms: (1) stridor; (2) oxygen saturation <90%; (3) chest indrawing; or (4) fast breathing.
<sup>2</sup> Computed for the total prevalence comparison. All prevalence ratios (PR) are unadjusted.
<sup>3</sup> Defined as receipt of vaccinations per national guidelines allowing for a one-month delay.
<sup>4</sup> Sanitation access was classified according to the JMP as unimproved (no facility, bush, field, flush toilets to elsewhere, pit latrines without slab floor, bucket toilets or hanging toilets/latrines), or improved (flush toilets to piped sewer systems, septic tanks or unknown location, ventilated improved pit latrines, pit latrines with slab floors, or composting toilets).
<sup>5</sup> Drinking water source was classified according to the WHO/UNICEF Joint Monitoring Programme for Water Supply, Sanitation and Hygiene (JMP) as unimproved (water from a river, dam, lake, pond, stream, canal, irrigation channel, or unprotected wells or springs, or improved (water piped into the dwelling, yard or plot, public taps or standpipes, tube wells or boreholes, protected dug wells or springs, rainwater collection, water from a tanker-truck, water from a cart with a small tank or drum, filtered or unfiltered water kiosks, bottled water, or sachet water).
<sup>6</sup> Defined as in Kotloff, Lancet GH, 2019. Moderate-to-severe diarrhea (MSD) defined as presenting to a health facility with diarrhea and severe or some dehydration (by WHO criteria), visible blood in stool, or inpatient admission. Less-severe-diarrhea (LSD) defined as presenting to a health facility without MSD.
<sup>7</sup> Defined as in PATH Vesikari Clinical Severity Scoring System Manual: Duration of diarrhea: 1-4 days (1 point), 5 days (2 points) ≥6 days (3 points); Max # of stool in 24 hour period: 1-3 (1 point), 4-5 (2 points), ≥6 (3 points); Duration of vomiting: 1 day (1 point), 2 days (2 points), ≥3 days (3 points); max # of vomiting episodes in 24 hour period: 1 (1 point), 2-4 (2 points), ≥5 (3 points); Axillary temperature 36-37.9°C (1 point), 38-0-38.4°C (2 points), ≥38.5°C (3 points); dehydration 1-5% (2 points), ≥6% (3 points); treatment: rehydration (1 point), hospitalization (2 points).
<sup>8</sup> Severe stunting: HAZ/LAZ < -3, Moderate stunting: -3 ≤ HAZ/LAZ < -2.

**Figure 2.**
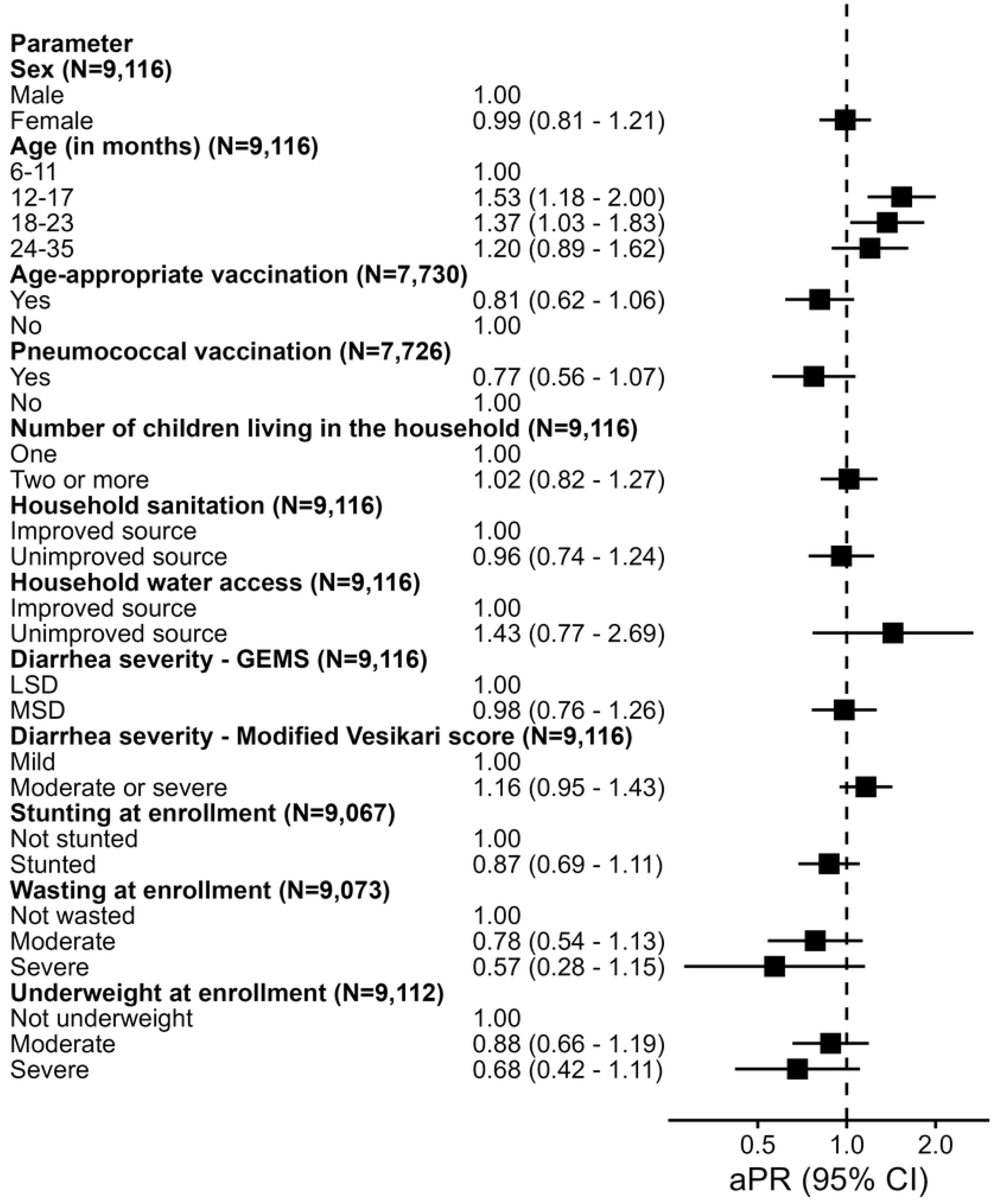
Prevalence of RTI by EFGH country site and age.

After adjusting for potential confounders, age was the only independent predictor of RTI during the diarrhea convalesce period: Children 12–17 months of age and those 18-23 months of age were approximately 1.5 times more likely to experience RTI during follow-up compared to children 6-11 months (adjusted prevalence ratio [aPR]: 1.53, 95% confidence interval [CI]: 1.18-2.00 and aPR: 1.37, CI: 1.03-1.83, respectively) (Figure 3). While statistically significant differences were observed prior to adjustment (Table 3), there was no association between RTI and receipt of all age-appropriate vaccinations after adjusting for xyz (aPR: 0.81, CI: 0.62-1.06) or pneumococcal vaccination (aPR: 0.77, CI: 0.56-1.07).

**Fig 3.**
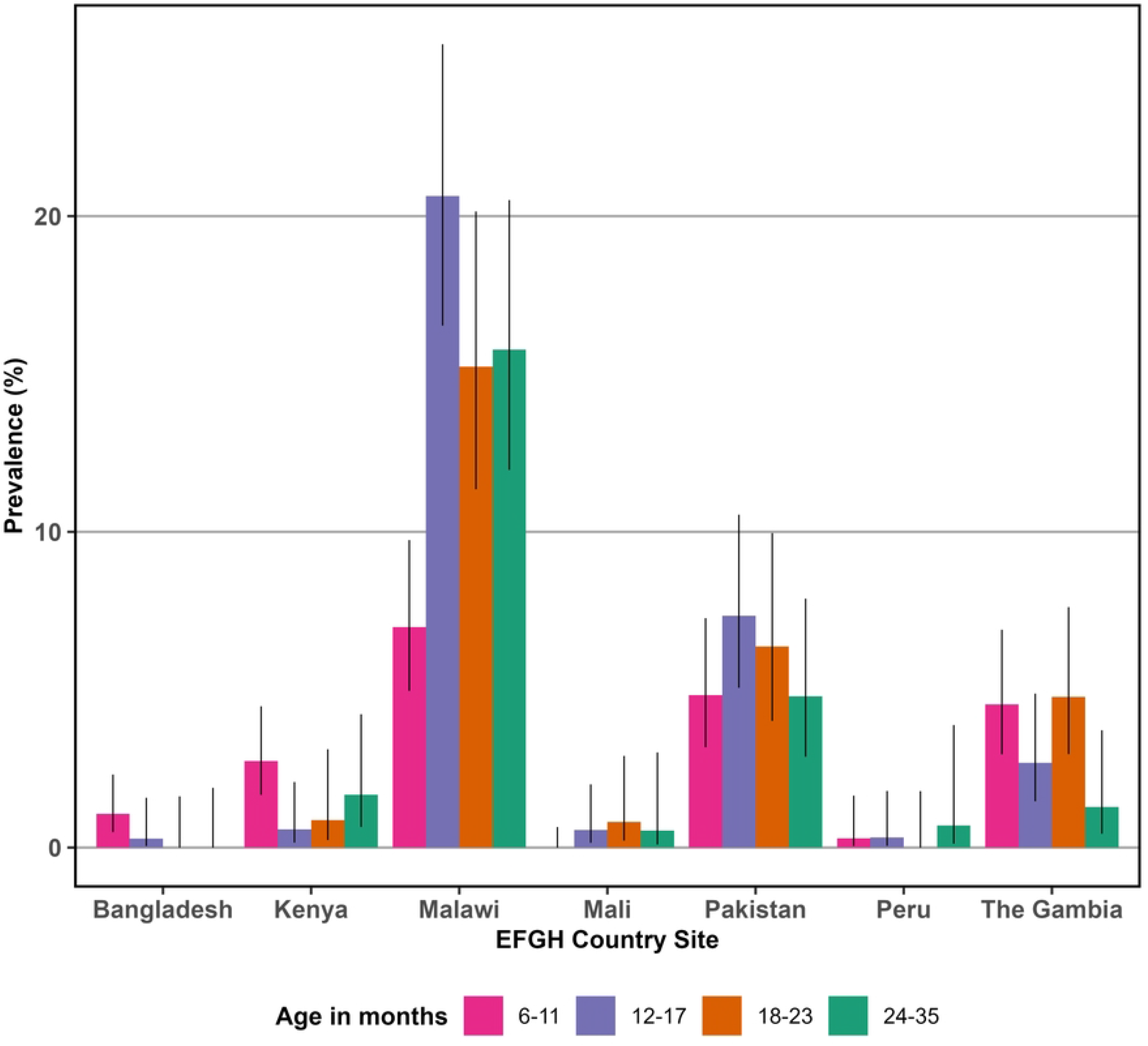
Correlates of RTI during follow-up.

## Discussion

We used EFGH-*Shigella* Surveillance Study data to estimate the period prevalence and risk factors of RTIs in the three months following MAD among children aged 6-35 months in Bangladesh, Pakistan, The Gambia, Malawi, Mali, Kenya and Peru. Our most important findings are that:

The observed proportion of diarrhea cases experiencing RTI during follow-up, with a notable concentration of cases happening in the first month, is concerning given the short follow-up period and likely dual burden of these two acute illnesses in young children. This finding suggests a temporal vulnerability in the immediate post-diarrheal recovery phase, possibly linked to compromised nutritional and immune status.(16)

Respiratory tract infections were more commonly observed among older children (12-24 months), a finding supported by previous literature indicating increased exposure to respiratory pathogens in children at this age.(17) This age-specific trend may also reflect waning maternal antibody protection, emphasizing the need for attention to vaccine timeliness and catch-up strategies during the second year of life.(18) This age group also coincides with a timeframe of high diarrhea burden, particularly diarrhea attributed to *Shigella*, which peaks in toddlers.(19)

The most frequently reported symptom of the RTI definition was cough (20.6%), with substantial variation between country sites (range 2.4% to 58.3%). This wide inter-site variability likely reflects differences in health-seeking behaviour and environmental exposure.(2) The demographic profile of the analysed cohort reflects a balanced representation across all seven countries. The predominance of children aged 12-17 months, with a median age of 15 months is consistent with the age window during which kids in LMICs are particularly susceptible to infectious diseases.(2,19)

Children without age-appropriate immunization were more likely to experience RTI (with the highest prevalence among those not receiving pneumococcal conjugate vaccine (PCV).). Multivariate analysis demonstrated that PCV receipt protected 27% of children against RTI, however this finding became less significant after adjusting for country site, age, and sex. PCV’s effectiveness in reducing all-cause and pneumococcal pneumonia has been demonstrated previously(20–22) and the lack of independent association in this study could be due to the fact that non-pneumococcal attributed respiratory infections were responsible for the RTI found during follow-up. We found inter-country variability in routine immunization coverage with Bangladesh attaining the highest coverage levels of age-appropriate EPI vaccine uptake (77.3%), while The Gambia recorded the lowest (27.4%). This disparity may reflect differences in health system performance, access barriers, or vaccine hesitancy.(23) Notably, PCV coverage was high overall (87.6%), in alignment with global efforts to scale up PCV introduction and access through initiatives like GAVI.(24)

Unlike previous studies, we did not find evidence of undernutrition increasing susceptibility to respiratory infection.(14,25) In fact, RTI appeared to be more common in children without wasting, although this was not significant after accounting for age, country site, and sex.

This analysis had several strengths, including its large sample size and representation from seven countries around the world. However, there were several limitations. The symptoms (cough or difficult breathing) of outcome of interest of RTI, were also caregiver-reported, and although useful in population-based research, it may be subject to recall or reporting bias. However, RTI was confirmed with presence of associated clinical findings in addition to reported symptoms by caregiver. Clinical confirmation of RTI through radiography or laboratory diagnostics would enhance specificity. Additionally, the relatively short follow-up period (three months) may underestimate the long-term impact of MAD episodes on respiratory health.

This analysis reinforces the interconnectedness of diarrheal and respiratory illnesses in early childhood and identifies key modifiable risk factors for post-diarrheal RTI. Strengthening immunization and improving sanitation are essential components of holistic child health strategies in LMICs. Future research should explore the biological mechanism linking enteric and respiratory infections and evaluate the impact of integrated interventions in reducing the compounding burden of pediatric infectious diseases.

## Acknowledgements

We are deeply appreciative of the contributions from the EFGH study teams across the 7 country sites (Bangladesh, Kenya, Malawi, Mali, Pakistan and The Gambia), and most importantly, the study participants and their families, whose involvement made this research possible. I also acknowledge the generous support of the Gates Foundation, and the excellent coordination efforts provided by the University of Washington. Special thanks to the Nyanja Health Research Institute in Malawi for their support and collaboration.

## Data Availability Statement

The EFGH statistical analysis plan (https://clinicaltrials.gov/study/NCT06047821) and study protocol (https://academic.oup.com/ofid/issue/11/Supplement_1) were made publicly available. The datasets were deidentified and anonymized and will be publicly available upon publication of the manuscript.

